# Implementing midwifery twinning partnerships: challenges and facilitators. A rapid evidence summary

**DOI:** 10.1101/2023.06.14.23291377

**Authors:** Deborah Edwards, Judit Csontos, Grace Thomas

## Abstract

Twinning is a partnership method that focuses on mutual transfer of knowledge and skills between two parties, including organisations, clinical practices, universities, or individual health professionals. In midwifery, twinning is a particularly important tool that can help countries with high maternal and infant morbidity and mortality rates to make connections with countries where sickness and death rates related to birth are lower and the role of midwives is better developed. The aim of this rapid evidence summary is to explore the literature for midwifery twinning initiatives and the facilitators and challenges of twinning partnerships.

Sixteen research reports and textual evidence were identified.

Facilitators of successful implementation of twinning initiatives include having a clear vision and mission statement along with investing time and promoting a co-creational approach. Reciprocity along with the building of personal relationships. Strong leadership, commitment, values, mutual respect and personal rapport between the projects. Clear communication plans, workshops, peer exchange visits alongside regular virtual contact. Building on existing relationships, previous experience of international and cross-cultural work and being prepared to overcome cultural differences. Having a local project team and careful; matching and selection of twins and having an adaptable personality. Having funding available.

Challenges include communication issues, cultural differences in communication, technological issues and economic considerations. Additionally misplaced expectations, such as difference in social expectations, or one twin partner expecting opportunities that are not agreed upon by the other poses challenges to the successful implementation of twinning initiatives.

## Wales Centre For Evidenced Based Care (WCEBC) Rapid Evidence Summary

### Implementing midwifery twinning partnerships: challenges and facilitators

#### EXECUTIVE SUMMARY

##### What is a Rapid Evidence Summary?

This Rapid Evidence Summary (RES) is designed to provide an interim evidence briefing to inform further work and provide early access to key findings. It is based on a limited search of key resources and no quality appraisal or evidence synthesis was conducted, and the summary should be interpreted with caution.

##### Who is this summary for?

World Health Organisation Collaborating Centre for Midwifery Development, Cardiff University, Wales, UK.

##### Key Findings

Sixteen research reports and textual evidence were identified.

*Extent of the evidence base*

∎ Types of included reports included descriptions of twinning initiatives with (n=2) or without a case study (n=7), a Delphi study (n=1), qualitative studies (n=2), mixed methods studies (n=2), a systematic review (n=1) and a literature review (n=1)
∎ Twinning partnerships focused on relationships between the following high income and low and middle income countries (North-South partnerships)
  ∘ The Netherlands - Sierra Leone (n=3) or Morocco (n=2);
  ∘ United Kingdom – Cambodia (n=1), Nepal (n=2), Rwanda (n=1), or Uganda (n=5);
  ∘ Canada – Tanzania (n=2);
  ∘ Japan – Mongolia (n=1);
  ∘ Switzerland – Mali (n=1);
  ∘ Australia – Papua New Guinea (n=1)

*Recency of the evidence base*

∎ The rapid evidence summary included evidence available up until January 2023, with reports published between 2013 and 2022.

*Facilitators*

- Reciprocity along with the building of personal relationships.
- Having a clear vision and mission statement along with investing time and promoting a co-creational approach.
- Strong leadership, commitment, values, mutual respect and personal rapport between the projects.
- Clear communication plans, workshops, peer exchange visits alongside regular virtual contact
- Building on existing relationships, previous experience of international and cross-cultural work and being prepared to overcome cultural differences.
- Local project team
- Careful; matching and selection of twins and having an adaptable personality.
- Funding

*Challenges*

∎ Communication issues between different training institutions, employers, and stakeholders.
∎ Cultural differences in communication, the meaning of a twin, and the lack of preparedness to bridge differences in culture.
∎ Technological issues, such as limited access to reliable internet, connectivity issues, different IT systems, and the lack of opportunities for in-person meeting.
∎ Monitoring and evaluating twinning initiatives can be problematic due to cultural differences in gathering data and the absence of indicators on what is considered success in a twinning partnership.
∎ Economic considerations, such as insufficient funding for travel or having a coordinator can be a barrier.
∎ Misplaced expectations, such as difference in social expectations, or one twin partner expecting opportunities that are not agreed upon by the other.
∎ Human resource issues including staff turnover or shortages, or overburdening twinning participants.

##### Next steps

∎ To conduct a systematic review focusing on the barriers and facilitators of twinning initiatives that could be broadened to include other healthcare professionals, such as nurses. The planned review could help inform the development of future twinning partnerships.

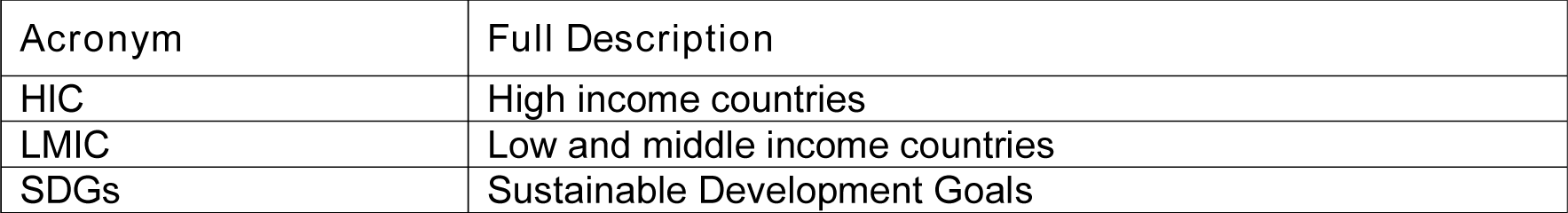
Abbreviations.

Abbreviations:
| Acronym | Full Description |
| --- | --- |
| HIC | High income countries |
| LMIC | Low and middle income countries |
| SDGs | Sustainable Development Goals |

## 1. CONTEXT / BACKGROUND

The United Nations adopted Sustainable Development Goals (SDGs) to help eradicate poverty while sustaining peace and freedom across the globe (United Nations 2015). The SDGs focus on 17 areas, which include ending poverty and hunger; achieving good health and well-being, quality education, clean water and sanitation, affordable and clean energy, decent work and economic growth for all, responsible consumption and production, peace and justice; ensuring gender equality and reducing inequalities; improving industry, innovation, and infrastructure; making cities sustainable; taking action on climate change; and preserving life in the water and on land (United Nations 2015). Finally, for these goals to become reality, global partnerships are required (United Nations 2015).

For healthcare professionals, achieving the goals of good health and wellbeing and gender equality are particularly important (Cadee 2020). As differences exist between healthcare systems and care provision around the world, collaboration between different countries could help support achieving these goals, while fulfilling the need for global partnership. Twinning is a partnership method that focuses on mutual transfer of knowledge and skills between two parties, including organisations, clinical practices, universities, or individual health professionals (Moyo and Bokosi 2014). In midwifery, twinning is a particularly important tool, as it can help countries with high morbidity and mortality rates among expectant mothers and infants to make connections with countries where sickness and death rates related to birth are lower and the role of midwives is better developed (Moyo and Bokosi 2014). Twinning can contain many activities, including on-site training, study tours, collaboration on technical initiatives, and training, information and technical exchanges (Moyo and Bokosi 2014). The benefits of twinning can be wide ranging, from capacity building to the exchange of best practices and networking, resulting in mutual gains for both parties involved (Moyo and Bokosi 2014). However, challenges to building a successful twinning partnership can also exist, including cultural differences and economic divide among others (Moyo and Bokosi 2014). For a twinning initiative to be successful, there is a need for high quality research to determine its effectiveness and the barriers and facilitators to its implementation and uptake (Cadee 2020). Therefore, the aim of this rapid evidence summary is to explore the literature for midwifery twinning initiatives and the barriers and facilitators of twinning partnerships.

## 2. RESEARCH QUESTION(S)

| Review question |  |
| --- | --- |
| What are the barriers and facilitators when implementing midwifery twinning initiatives |  |
| Participants | Midwives and midwifery students |
| Concept | Twinning partnerships |
| Context | North-South partnerships* |
| Other Study Considerations |  |
| English language publications, preprints and organisational reports |  |
| Date restrictions – None |  |

\*North-South partnerships: Partnerships between midwifery organisations and midwives in LMIC and HIC

## 3. SUMMARY OF THE EVIDENCE BASE

### 3.1. Type and amount of evidence available

Sixteen research reports and textual evidence were included: seven descriptions of twinning initiatives (Adhikari and Nsubuga 2017; Cadee et al. 2013; Kemp et al. 2018a, Kemp et al. 2018b; Kemp et al. 2018c; Royal College of Midwives 2015; Sandwell et al. 2018); two descriptions of twinning initiative and case studies of midwives’ experiences (Ireland et al.

2015; Moyo and Bokosi 2014); one Delphi study (Cadee et al. 2018); two qualitative studies (Cadee et al. 2021a, Cadee et al.2021b); two mixed method studies (Taniguchi et al. 2022; Voluntary Services Overseas 2022); one systematic review (Dawson et al. 2014); and one literature review (Cadee et al. 2016).

- An evaluation of the Twin2Twin method which seeks to enhance the platform of exchange at an organisational level with the introduction of a personal exchange between individual midwives. This project involved the mutual exchange of knowledge and skills for midwives in Sierra Leone and the Netherlands (Cadee et al. 2013).
- A concept analysis of twinning in healthcare and the development of an operational definition of twinning from the international literature (Cadee et al. 2016).
- A Delphi study with 56 midwives from 19 countries to gain consensus for critical success factors associated with twinning in midwifery (Cadee et al. 2018).
- A qualitative study that explored the contribution of one-to-one relationships between twins to twinning projects, in Morocco and the Netherlands and Sierra Leone and the Netherlands (Cadee et al. 2021a).
- A qualitative study that explored how the professional growth of midwives can be enhanced through twinning collaborations for midwives in Morocco, Sierra Leone and the Netherlands (Cadee et al. 2021b).
- A systematic review that explored the collaborative approaches undertaken to build midwifery education regulation and professional association in low-income countries (partnerships between midwifery organisations and midwives in low and middle income countries (LMIC) described as South–South partnerships or between LMIC and high income countries described as North–South partnerships) (Dawson et al. 2014).
- Two papers that cover different aspects of the Global Midwifery Twinning Project which was a three-year multi-country partnership running from 2012 to 2015 to strengthen health systems through health service skills transfer and capacity development between the UK and three partner organisations (Cambodia, Nepal and Uganda).
  ∘ A paper that describes the importance of collaboration between established national midwifery organisations and newly established ones as part of the Global Midwifery Twinning Project and highlights the experience of a UK midwifery volunteer in Nepal (Ireland et al. 2015).
  ∘ The final evaluation report for the project (Royal College of Midwives 2015).
- Four papers that cover different aspects of the MOMENTUM project which is a twinning initiative between UK and Uganda.
  ∘ A paper that describes the development of a national standard for midwifery mentorship in Uganda and explores twinning partnerships as part of this (Kemp et al. 2018a).
  ∘ A paper that describes the development of the MOMENTUM initiative, a 20-month midwifery twinning project aiming to develop a model of mentorship for Ugandan midwifery students (Kemp et al. 2018b).
  ∘ A paper describing a practice development workstream, a twinning project that used work-based learning and appreciative inquiry, embedded in an action research approach, to facilitate practice (Kemp et al.2018c).
  ∘ The final project evaluation report (Adhikari and Nsubuga 2017)
- A mixed methods study that evaluated the outcomes of the organisational strengthening of the Mongolian Midwives Association through the twinning project with Japanese Midwives Association (Taniguchi et al. 2022).
- A report of a mixed method evaluation that assesses the impact of the twinning partnership between Nyagatare District Hospital (NDH) in Rwanda and Lewisham Sexual Health Services (LSHS) within Lewisham and Greenwich NHS Trust (LGT) in the UK (Voluntary Services Overseas 2022).
- A paper that describes a successful twinning relationship between midwifery association in Canada and Tanzania, to analyse how and why it worked, and what benefits it provided (Sandwell et al. 2018).
- The operational manual of the International Confederation of Midwives on twinning as a tool for strengthening midwives associations (Moyo and Bokosi 2014).
- Specific twinning initiatives were reported between the following LMICS and HICs:
  ∘ Sierra Leone and the Netherlands (Cadee et al. 2013; Cadee et al. 2021a; Moyo and Bokosi 2014)
  ∘ Morocco and the Netherlands (Cadee et al. 2021a; Cadee et al. 2021b)
  ∘ Nepal and the UK (Ireland et al. 2015; Royal College of Midwives 2015)
  ∘ Uganda and UK (Adhikari and Nsubuga 2017; Kemp et al. 2018a, Kemp et al. 2018b; Kemp et al. 2018c; Royal College of Midwives 2015)
  ∘ Rwanda and UK (Voluntary Services Overseas 2022)
  ∘ Cambodia and UK (Royal College of Midwives 2015)
  ∘ Tanzania and Canada (Moyo and Bokosi 2014; Sandwell et al. 2018)
  ∘ Mongolia and Japan (Taniguchi et al. 2022)
  ∘ Mali and Switzerland (Moyo and Bokosi 2014)
  ∘ Papua New Guinea and Australia (Moyo and Bokosi 2014)

### 3.2. Key Findings

#### 3.2.1. Positive outcomes of twinning initiatives

- The systematic review conducted by Dawson et al. 2014 was unable to locate an example of twinning in international midwifery contexts (North to South).
- MOMENTUM - a 20 month midwifery twinning project aiming to develop a model of mentorship for Ugandan midwifery students was successful at improving midwifery students in knowledge, skills, attitude and self-reported levels of confidence and competence to provide quality midwifery care (Kemp et al. 2018b).
- The concept analysis of the literature resulted in four main attributes of twinning in healthcare. Most important was reciprocity along with the building of personal relationships, being a dynamic process and that twinning is between two named organisations across different cultures (Cadee et al. 2016).
- The definition that was developed from this was that *‘Twinning is a cross-cultural, reciprocal process where two groups of people work together to achieve joint goals’.* (Cadee et al. 2016, p. 8.)
- Positive outcomes were demonstrated as a result of the twinning project between the Japanese Midwives Association and Mongolian Midwives Association (MMA) when organisational strengthening was evaluated through the Member Association Capacity Assessment Tool. Positive opinions and impressions of the twinning project were also elicited through qualitative interviews (Taniguchi et al.2022).
- The Global Midwifery Twinning Project (Royal College of Midwives 2015) had beneficial effects on midwifery practice, education and regulation in the three partner countries (Cambodia, Nepal and Uganda).

#### 3.2.2. Facilitators of successful implementation of twinning initiatives

- Investing time at the beginning of the partnership to develop relationships (Voluntary Services Overseas, 2022).
- Promoting a co-creational approach, where the twin institutions co-design the scoping and work planning phases of the project (Voluntary Services Overseas, 2022).
- Conducting a thorough risk assessment at the start to mitigate the risks associated with staff turnover (Voluntary Services Overseas 2022).
- Establishing a twinning partnership that builds on existing relationships (Voluntary Services Overseas, 2022).
- Strong leadership, commitment, mutual respect and personal rapport between the project leads in each twin institution (Sandwell et al. 2018; Voluntary Services Overseas, 2022).
- Regular virtual contact between visits (Kemp et al. 2018a).
- Clear and agreed communication plans (Cadee et al. 2018; Kemp et al. 2018a).
- Commitment and values (Cadee et al. 2018).
- Making equity explicit in twinning may contribute towards the agency of midwives to take on their identified key role in sexual and reproductive healthcare (Cadee et al. 2018).
- Building trusting relationships (Cadee et al. 2021a).
- Being prepared to overcome cultural differences (Cadee et al. 2021b).
- The development of common midwifery goals (Cadee et al. 2021b).
- Having an adaptable personality (Cadee et al. 2021b).
- A successful twinning partnership is a reciprocal and equitable environment (Cadee et al. 2021b; Voluntary Services Overseas 2022).
- Formation of a social media network via Whatsapp (Kemp et al. 2018a).
- Careful matching and selection of twins (Kemp et al. 2018a).
- A local project team who can make frequent visits to project sites and act as trouble shooters when problems arise (Kemp et al. 2018a).
- Workshops where all participants can reflect, observe and plan (Kemp et al. 2018a).
- Peer exchange visits (Kemp et al. 2018a).
- Repeated placements of short term international heath volunteers within the context of a long term partnership (Kemp et al. 2018a).
- Having a clear vision and mission statements (Sandwell et al. 2018).
- Being familiar with international midwifery (Sandwell et al. 2018).
- International global health climate and trends in donor priorities (Sandwell et al. 2018).
- Previous experience with international and cross-cultural work (Sandwell et al. 2018).
- Model of care which emphasises partnership throughout midwives work (Sandwell et al. 2018).
- Funding, both for the initial twinning relationship and for the shared project (Sandwell et al. 2018)

#### 3.2.3. Barriers to successful implementation of twinning initiatives

- Communication issues (Adhikari and Nsubuga 2017; Adhikari and Nsubuga 2017; Voluntary Services Overseas 2022).
  ∘ For example between different nurse /midwife training institutions and workforce employing institutions, between stakeholders (Adhikari and Nsubuga 2017; Adhikari and Nsubuga 2017).
- Cultural differences (Cadee et al. 2013; 2021a, Ireland et al. 2015; Kemp et al. 2018a; Moyo and Bokosi 2014)
  ∘ In communication (Cadee et al. 2013; Kemp et al, 2018a).
  ∘ What it means to be a twin (Cadee et al.2021a).
  ∘ Midwives were unprepared to bridge cultural differences (Cadee et al.2021b).
- Technological issues (Adhikari and Nsubuga 2017; Cadee et al. 2013; Kemp et al. 2018a, Moyo and Bokosi 2014; Sandwell et al. 2018; Voluntary Services Overseas 2022).
  ∘ Limited access to reliable internet, telecommunication connectivity, electric power grid and poor computer skills (Cadee et al. 2013; Kemp et al. 2018a, Moyo and Bokosi 2014). Sandwell et al. 2018).
  ∘ Maintaining the partnership was more difficult when in-person meetings were not possible (Sandwell et al. 2018).
  ∘ Connectivity issues particularly through online communication channels such as Microsoft Teams or Zoom (Whatsapp is easier) Voluntary Services Overseas 2022).
  ∘ Different, uncoordinated IT systems across UK hospitals (Voluntary Services Overseas 2022).
- Time zones especially to organise planned meetings (Kemp et al. 2018a Sandwell et al. 2018).
- The role and position of women in society (Ireland et al. 2015).
- Personality and character differences between individual midwife twin pairs (Cadee et al. 2013; Cadee et al.2021a; Sandwell et al. 2018).
- Issues around coordination and facilitation such as attitudes, professionalism and clarity about process and expectations (Cadee et al. 2013).
- Midwifery is not recognised as a separate profession (Ireland et al. 2015).
- Busy schedules and limited time and energy and workload issues alongside competing responsibilities (Ireland et al. 2015; Kemp et al. 2018; Sandwell et al. 2018).
- Monitoring and evaluation concerns (Cadee et al. 2013; Voluntary Services Overseas 2022).
  ∘ Formal evaluation is challenging due to cultural differences in gathering information (storytelling as opposed to written evaluations) (Cadee et al. 2013).
  ∘ Insufficient focus on monitoring and evaluation and specifically the absence of indicators to determine what constitutes ‘success’ for the twinning project
- Economic considerations
  ∘ Midwives being able to fund themselves to travel and carryout activities (Moyo and Bokosi 2014)
  ∘ Lacking the funding for a coordinator to organise meetings between practicing midwives in both countries who were too busy to self-coordinate (Sandwell et al. 2018).
- Misplaced expectations (Adhikari and Nsubuga 2017; Moyo and Bokosi 2014).
  ∘ Minds and social expectations (Moyo and Bokosi 2014).
  ∘ For example, one of the twinning partners may expect the Twinning relationship to provide travel opportunities, while the other may see it as an opportunity to gain exposure through practicing in their Twinning partner’s country (Moyo and Bokosi 2014).
- Difficulty in operationalising the equal footing concept due to availability or lack of resources especially when one of the twin countries is an ex-colony (due to historical issues and perceptions) (Moyo and Bokosi 2014).
- Culture of giving and dependency (donor and recipient cultures) - the issue of one twin partner looking to its twin to continually give due to a culture of dependency (Moyo and Bokosi 2014).
- Difficulty in initiating the relationship effectively – not dealing with identified challenges in the set-up phase (Moyo and Bokosi 2014).
- Human resource issues (Adhikari and Nsubuga 2017; Voluntary Services Overseas 2022)
  ∘ Staff turnover (Voluntary Services Overseas 2022)
  ∘ Lack of human resources and shortage of staff (Voluntary Services Overseas 2022).
  ∘ Overburden of human resources – seen as an additional task for midwives to give support to midwifery students (Adhikari and Nsubuga 2017).
- Sustainability (Adhikari and Nsubuga 2017;Voluntary Services Overseas 2022)
- Future uncertainty and anxiety (Adhikari and Nsubuga 2017).
- Reliance on VSO funding and support - - to date, the twinning project has been heavily reliant on VSO funding, brokering and support; while this is expected in the short term, it is clearly not sustainable longer term (Voluntary Services Overseas 2022).

### 3.3. Areas of uncertainty

- The majority of included reports were descriptions of twinning initiatives indicating a lack of research in the field, particularly quantitative research.
- There is inconsistency in the use of the term twinning in the wider literature (Cadee et al. 2016), with twinning often classified as international collaboration or exchange. Thus, it is possible that more literature on twinning is available.
- There is a lack of clarity in how the process of twinning and its related contributions to midwifery and maternal health improvements are assessed, particularly in capacity building projects (Dawson et al. 2014). This could influence the methodological quality of studies, and the certainty in research findings and the available evidence.

## 4. NEXT STEPS

- A qualitative or mixed methods (including textual evidence) systematic review focusing on the barriers and facilitators of twinning initiatives that could be broadened to include other healthcare professionals, such as nurses.

## Data Availability

All data produced in the present work are contained in the manuscript

## 6. RAPID EVIDENCE SUMMARY METHODS

Five databases were searched (Cinahl, Embase, Global Health, Medline Ovid Emcare in December 2022 and the search strategies are provided in Appendix I. Search hits were screened for relevance by two reviewers. No formal quality assessment was conducted. Citation, recency, evidence type and key findings were tabulated for all relevant primary and secondary research identified in this process by one reviewer and checked for accuracy by a second.

## 7. EVIDENCE

A more detailed summary of included evidence can be found in Table 2.

**Table 1.** Summary of review evidence identified

| **Evidence type** | **Total identified** | **Comments** |
| --- | --- | --- |
| Description of twinning initiative | 7 |  |
| Description of twinning initiative and case study | 2 |  |
| Primary Studies | 5 | 1 Delphi study<br>2 Qualitative<br>2 Mixed methods |
| Literature review | 1 |  |
| Systematic reviews (SRs) | 1 |  |

**Table 2:**
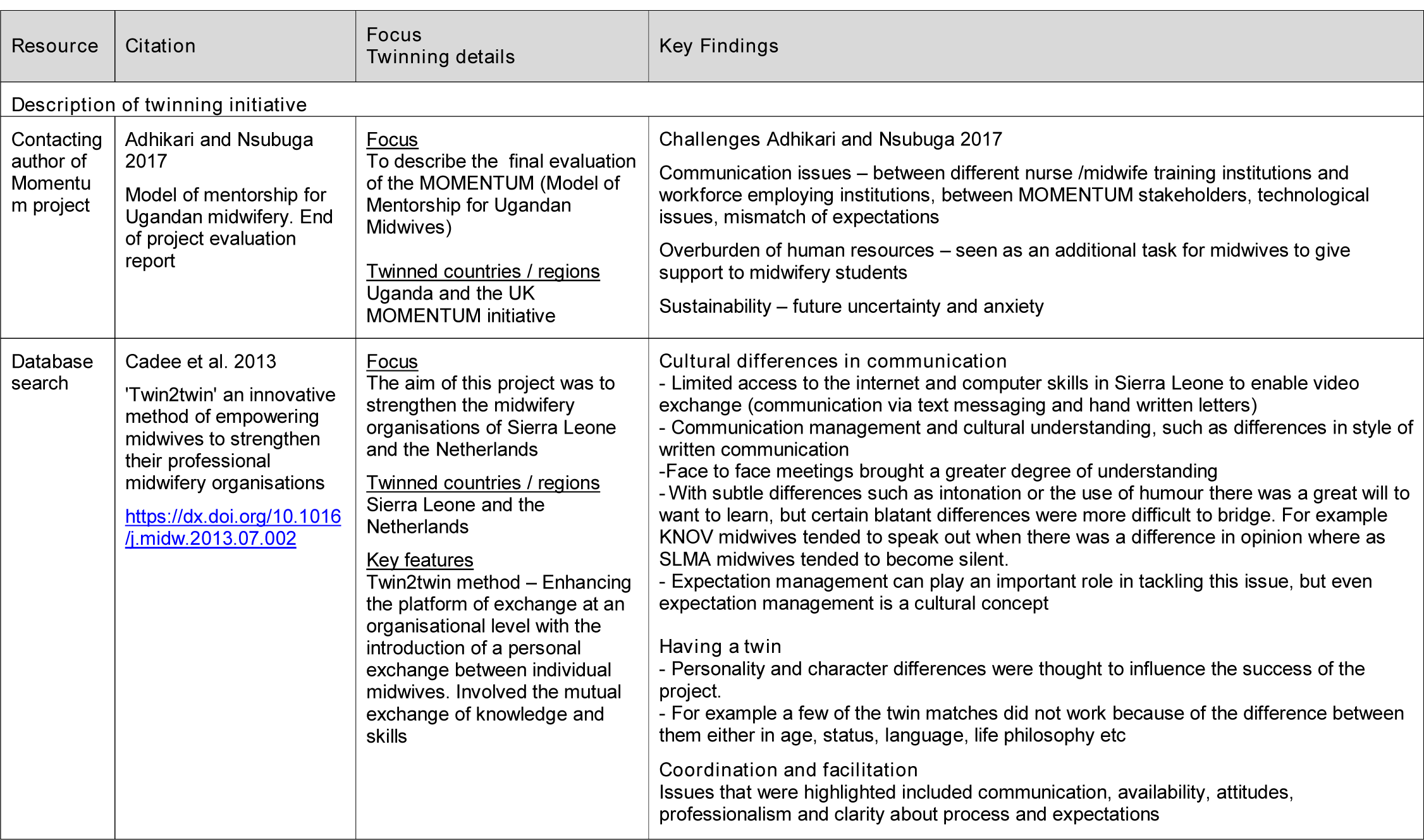
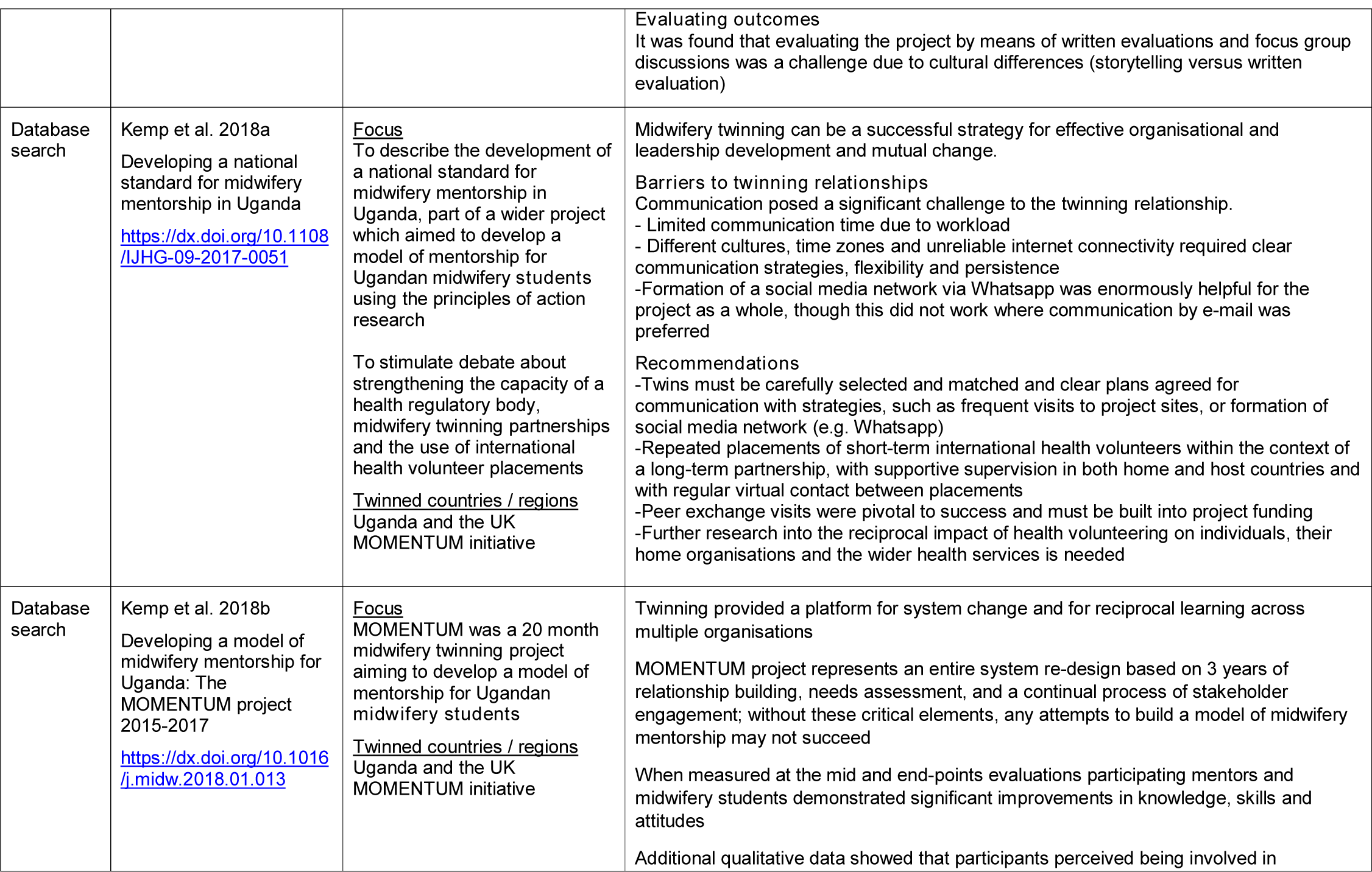
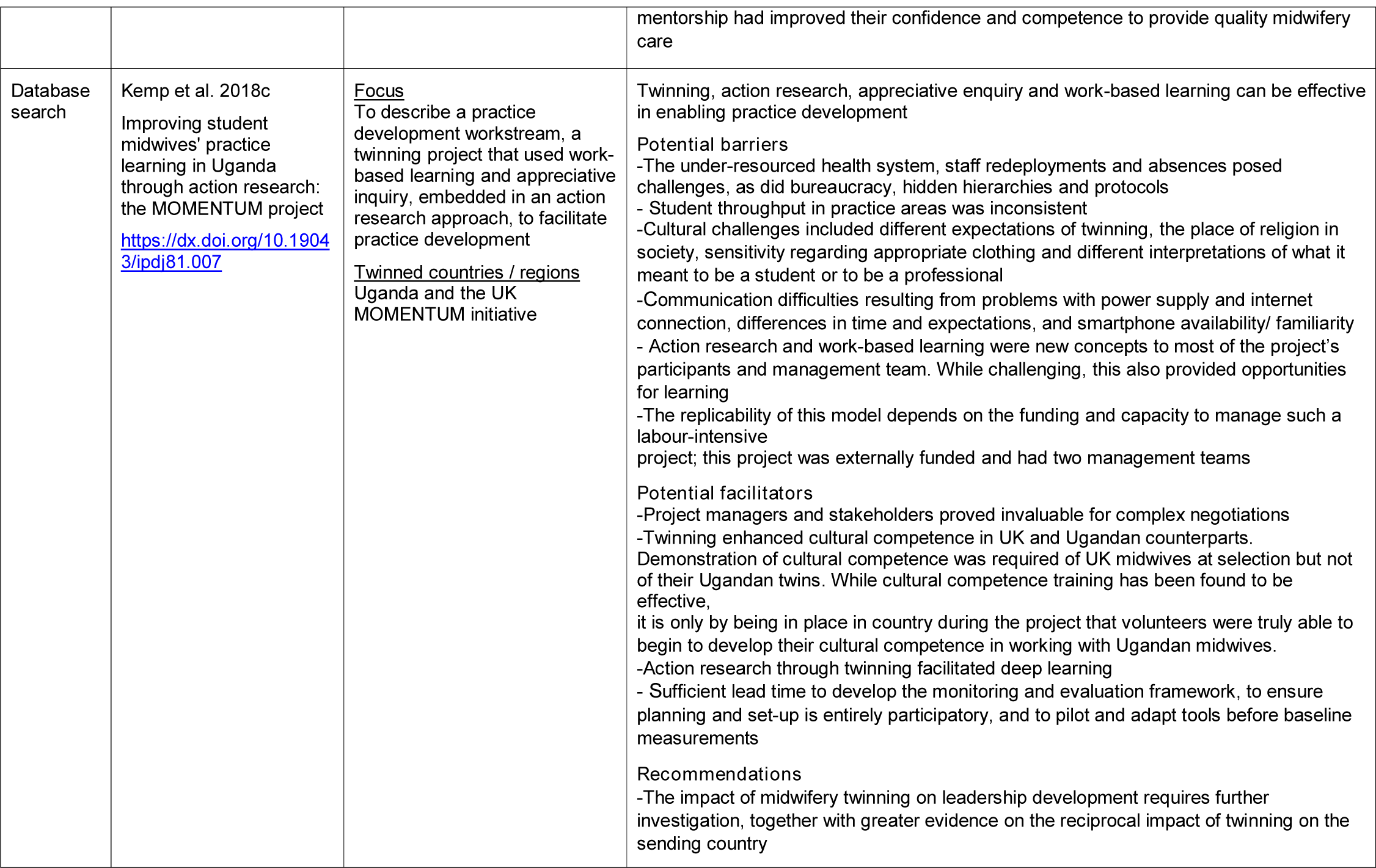
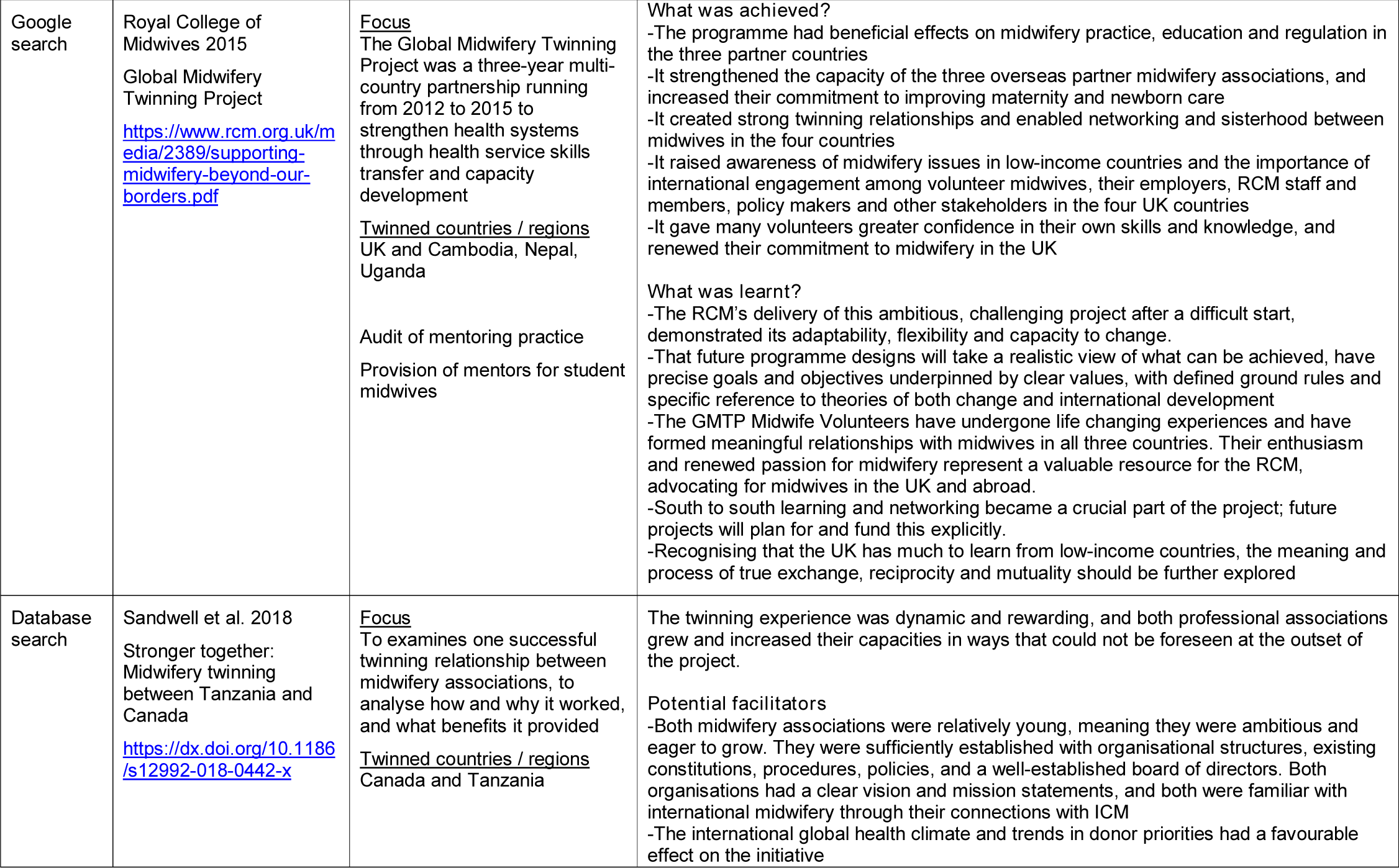
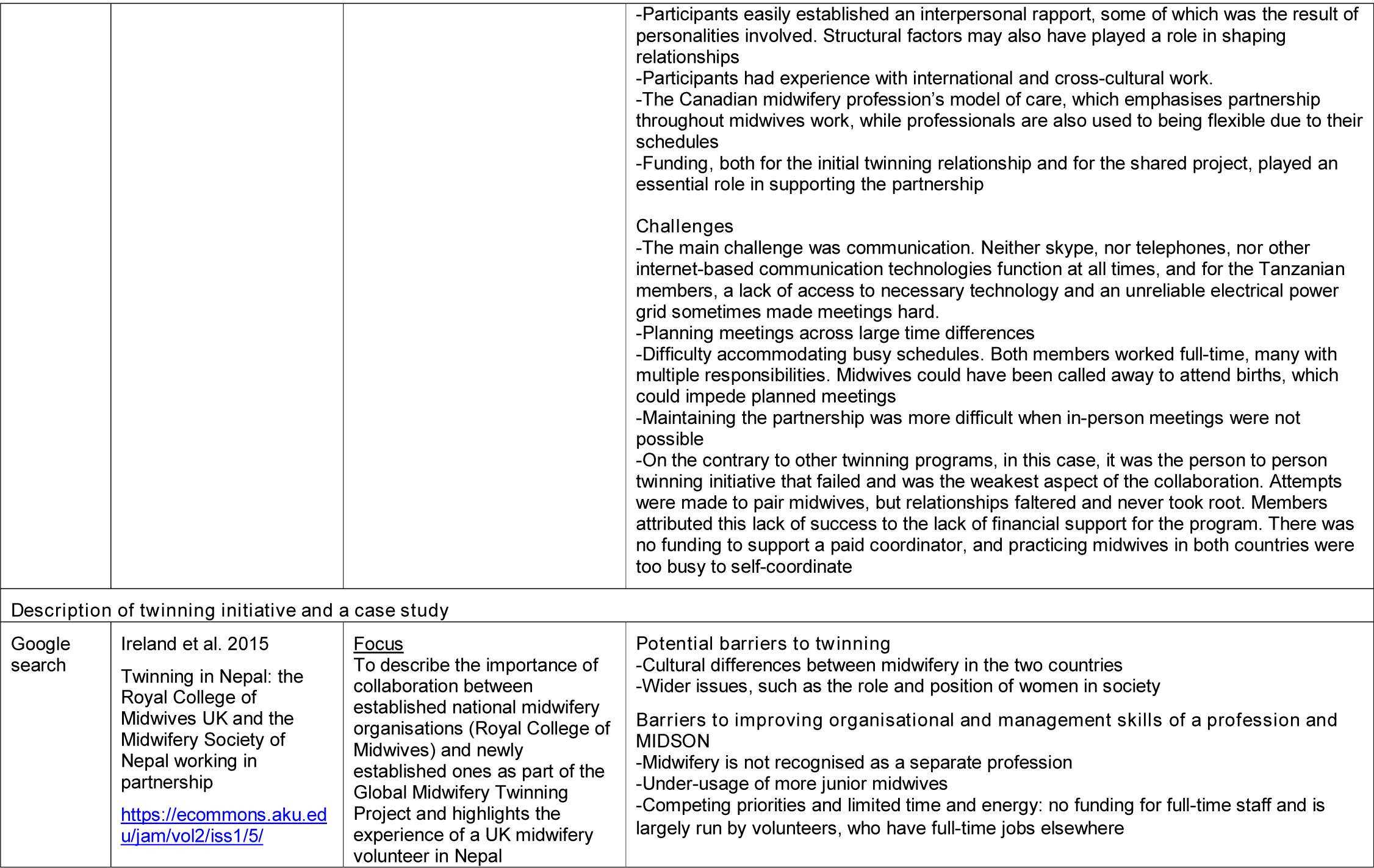
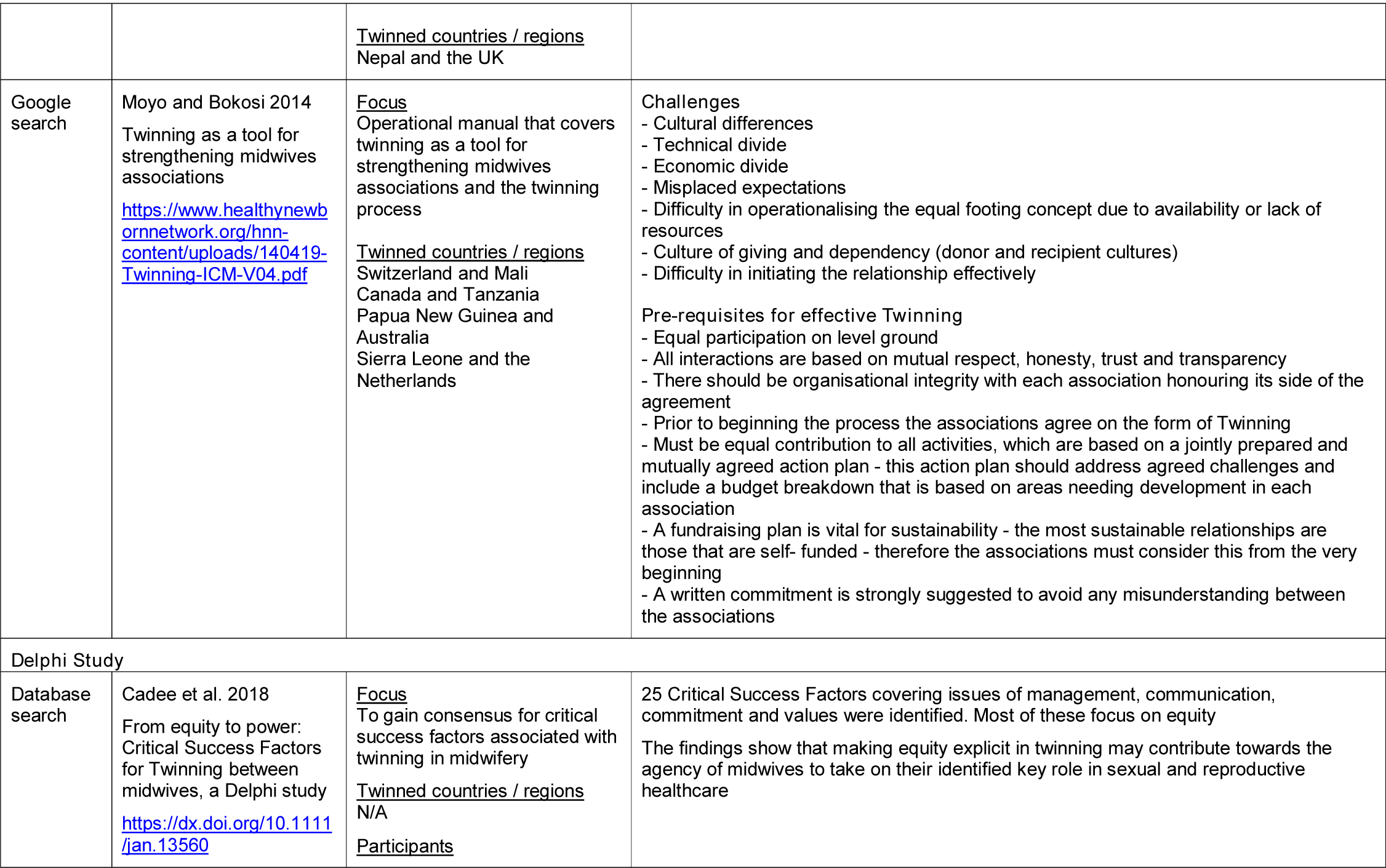
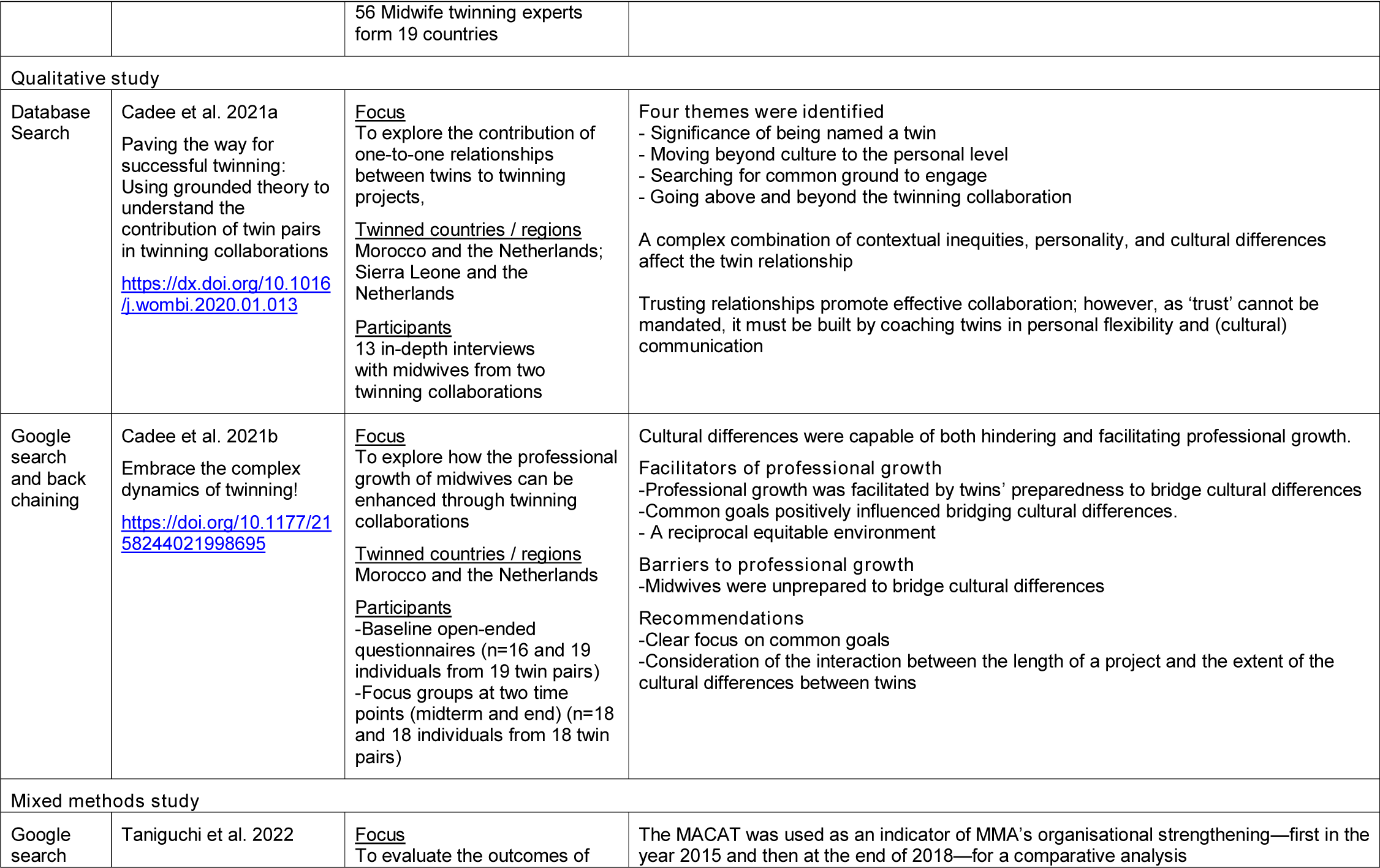
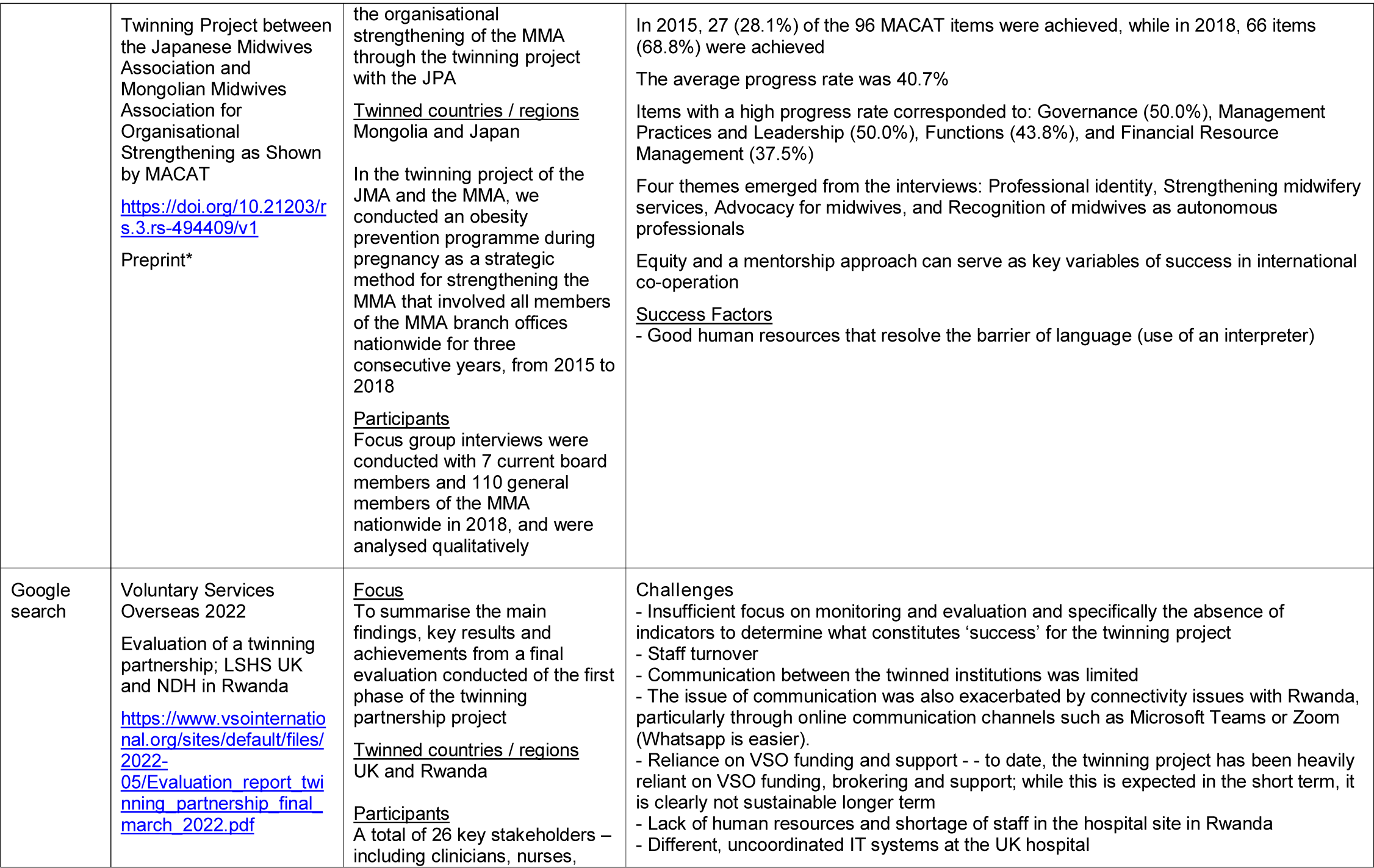
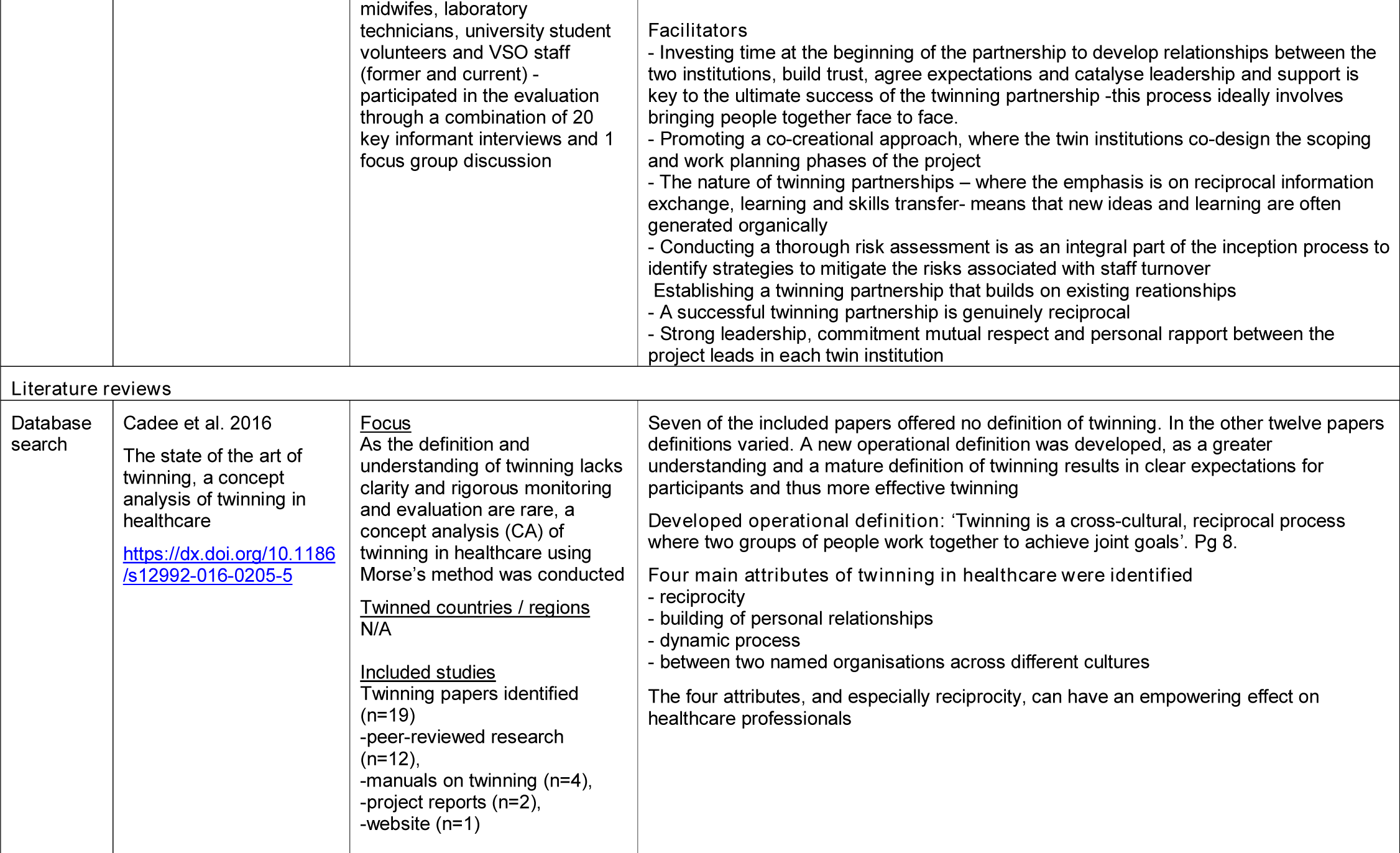
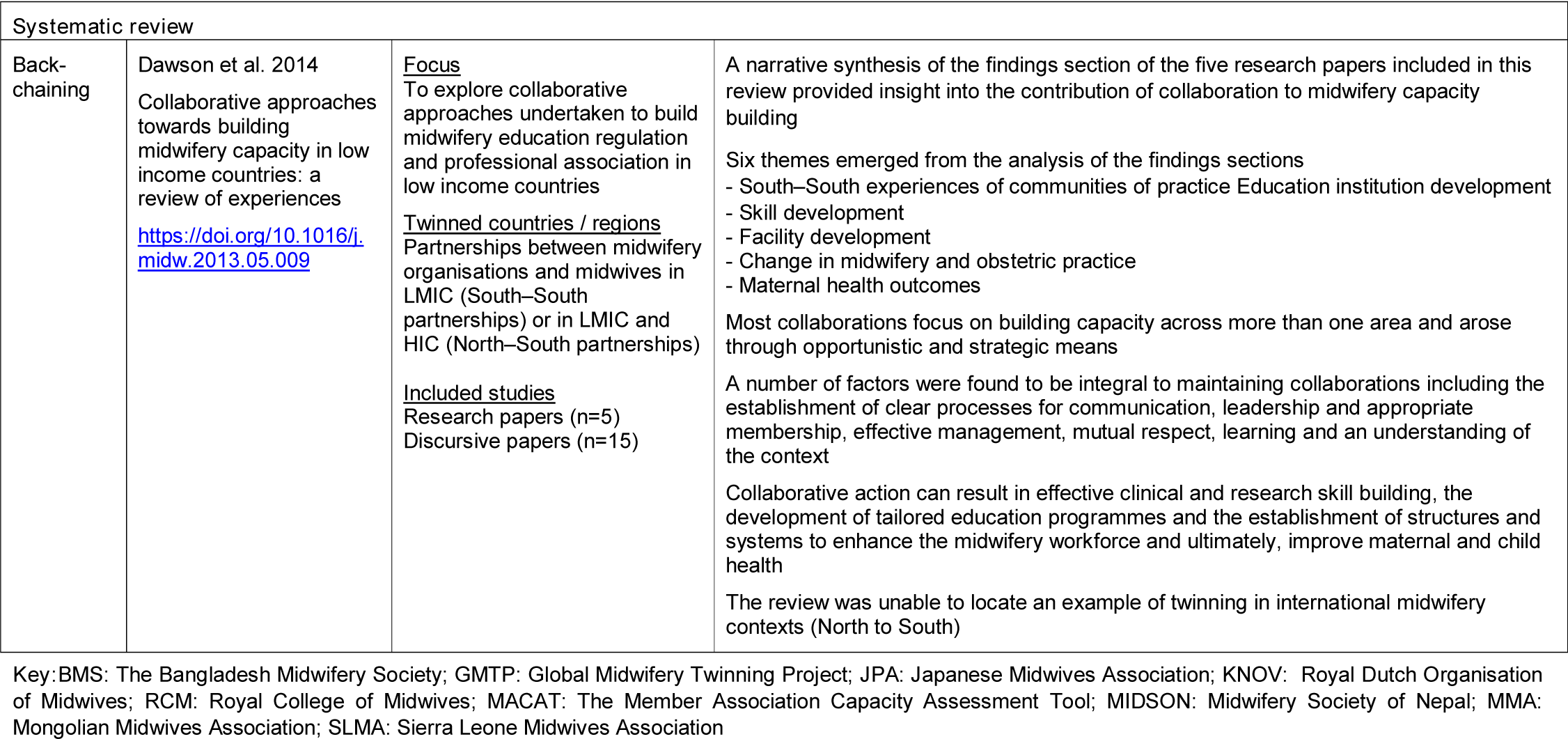
Summary of included evidence

## 8. ABOUT THE WALES CENTRE FOR EVIDENCE BASED CARE (WCEBC)

The WCEBC promotes evidence-based practice through the development and evaluation of internationally excellent systems for evidence appraisal, translation and utilisation.

We operate with a core team in the School of Healthcare Sciences, Cardiff University, and are led by Dr Deborah Edwards and Dr Clare Bennett.

We work closely with the following organisations: JBI, European Cancer Organisation, World Health Organisation Collaborating Centre for Midwifery Development and the Wales COVID-19 Evidence Centre.

**Co-Directors:**

Dr Deborah Edwards

Dr Clare Bennett

**Website:** https://www.cardiff.ac.uk/research/explore/research-units/wales-centre-for-evidence-based-care

## 9. APPENDIX

### 9.1. Search strategies

**Database: Embase <1974 to 2022 December 15>**

1. (midwif* or midwiv* or nurs* or doctor* or physician*).ti,ab. (1324447)
2. (health* adj2 (worker* or professional* or personnel or workforce)).ti,ab. (261325)
3. 1 or 2 (1511369)
4. (twinning or twinned or twin2twin).ti,ab. (4573)
5. (program* or collaborat* or project* or partner* or concept* or relation* or model* or capacity).ti,ab. (9093501)
6. 3 and 4 and 5 (148)
7. limit 6 to english language (147)
8. from 7 keep 1-147 (147)

**Database: Ovid MEDLINE(R) ALL <1946 to December 15, 2022>**

1. (midwif* or midwiv* or nurs* or doctor* or physician*).ti,ab. (1036576)
2. (health* adj2 (worker* or professional* or personnel or workforce)).ti,ab. (202708)
3. 1 or 2 (1185122)
4. (twinning or twinned or twin2twin).ti,ab. (4864)
5. (program* or collaborat* or project* or partner* or concept* or relation* or model* or capacity).ti,ab. (7289910)
6. 3 and 4 and 5 (59)
7. limit 6 to english language (59)

**Database: Global Health <1910 to 2022 Week 50>**

1. (midwif* or midwiv* or nurs* or doctor* or physician*).ti,ab. (122851)
2. (health* adj2 (worker* or professional* or personnel or workforce)).ti,ab. (64087)
3. 1 or 2 (173311)
4. (twinning or twinned or twin2twin).ti,ab. (305)
5. (program* or collaborat* or project* or partner* or concept* or relation* or model* or capacity).ti,ab. (1252341)
6. 3 and 4 and 5 (21)
7. limit 6 to english language (21)

**Database: Ovid Emcare <1995 to 2022 Week 48>**

1. (midwif* or midwiv* or nurs* or doctor* or physician*).ti,ab. (538725)
2. (health* adj2 (worker* or professional* or personnel or workforce)).ti,ab. (129703)
3. 1 or 2 (631009)
4. (twinning or twinned or twin2twin).ti,ab. (703)
5. (program* or collaborat* or project* or partner* or concept* or relation* or model* or capacity).ti,ab. (2264499)
6. 3 and 4 and 5 (28)
7. limit 6 to english language (28)

**Database: CINAHL: <inception to 16 December 2022)**

S1 (TI (midwif* or midwiv* or nurs* or doctor* or physician*)) OR (AB (midwif* or midwiv* or nurs* or doctor* or physician*)) (837,516)

S2 (TI (health* N2 (worker* or professional* or personnel or workforce)) OR (AB (health* N2 (worker* or professional* or personnel or workforce)) (131,522)

S3 TI (twinning or twinned or twin2twin) OR AB (twinning or twinned or twin2twin) (437)

S4 (TI (program* or collaborat* or project* or partner* or concept* or relation* or model* or capacity)) OR (AB (program* or collaborat* or project* or partner* or concept* or relation* or model* or capacity)) 1,681,866

S5 S1 OR S2 (928,527) S6 S3 AND S4 AND S5 (36)

## REFERENCES

1. Adhikari, R. and Nsubuga, F. 2017. Model of Mentorship for Ugandan Midwifery - MOMENTUM. End of project evaluation report. Royal College of Midwives, London.

2. Cadee, F. 2020. Twinning, a promising dynamic process to strengthen the agency of midwives. Maastricht University. Available from: https://cris.maastrichtuniversity.nl/ws/portalfiles/portal/52766210/s6807.pdf. Accessed 5th January 2023.

3. Cadee, F., Nieuwenhuijze, M. J., Lagro-Janssen, A. L. M. and De Vries, R. 2016. The state of the art of twinning, a concept analysis of twinning in healthcare. Globalization and Health 12(1). https://dx.doi.org/10.1186/s12992-016-0205-5

4. Cadee, F., Nieuwenhuijze, M. J., Lagro-Janssen, A. L. M. and de Vries, R. 2018. From equity to power: Critical Success Factors for Twinning between midwives, a Delphi study. Journal of Advanced Nursing 74(7): 1573–1582. https://dx.doi.org/10.1111/jan.13560

5. Cadee, F., Nieuwenhuijze, M. J., Lagro-Janssen, A. L. M. and de Vries, R. 2021. Paving the way for successful twinning:: Using grounded theory to understand the contribution of twin pairs in twinning collaborations. Women and Birth 34(1): 14–21. https://dx.doi.org/10.1016/j.wombi.2020.01.013

6. Cadee, F., Perdok, H., Sam, B., de Geus, M. and Kweekel, L. 2013. ‘Twin2twin’ an innovative method of empowering midwives to strengthen their professional midwifery organisations. Midwifery 29(10):1145–1150. https://dx.doi.org/10.1016/j.midw.2013.07.002

7. Cadée, F. M., Nieuwenhuijze, M. J., Lagro-Janssen, A. L. M. and de Vries, R. 2021. Embrace the complex dynamics of twinning!. SAGE Open 11(1). https://doi.org/10.1177/2158244021998695

8. Dawson, A., Brodie, P., Copeland, F., Rumsey, M. and Homer, C. 2014. Collaborative approaches towards building midwifery capacity in low income countries: a review of experiences. Midwifery 30(4):391–402. https://doi.org/10.1016/j.midw.2013.05.009

9. Ireland, J., van Teijlingen, E. and Kemp, J. 2015. Twinning in Nepal: the Royal College of Midwives UK and the Midwifery Society of Nepal working in partnership. Journal of Asian Midwives 2(1):26–33. https://ecommons.aku.edu/jam/vol2/iss1/5/

10. Kemp, J., Bannon, E. M., Mwanja, M. M. and Tebuseeke, D. 2018. Developing a national standard for midwifery mentorship in Uganda. International Journal of Health Governance 23(1):81–94. https://dx.doi.org/10.1108/IJHG-09-2017-0051

11. Kemp, J., Shaw, E. and Musoke, M. G. 2018. Developing a model of midwifery mentorship for Uganda: The MOMENTUM project 2015-2017. Midwifery 59, 27–129. https://dx.doi.org/10.1016/j.midw.2018.01.013

12. Kemp, J., Shaw, E., Nanjego, S. and Mondeh, K. 2018. Improving student midwives’ practice learning in Uganda through action research: the MOMENTUM project. International Practice Development Journal 8(1):1–23. https://dx.doi.org/10.19043/ipdj81.007

13. Moyo, N. T. and Bokosi, M. A. 2014. Twinning as a tool for strengthening Midwives Associations. The Hague: Available from: https://www.healthynewbornnetwork.org/hnn-content/uploads/140419-Twinning-ICM-V04.pdf. Accessed 5th January 2023.

14. Voluntary Services Overseas 2022. Evaluation of a twinning partnership; LSHS in UK and NDH in Rwanda. Available from: https://www.vsointernational.org/sites/default/files/2022-05/Evaluation_report_twinning_partnership_final_march_2022.pdf. Accessed 5th January 2023.

15. Royal College of Midwives 2015. Global Midwifery Twinning Project: Supporting Midwifery Beyond Our Borders. Royal College of Midwives, London. Available from: https://www.rcm.org.uk/media/2389/supporting-midwifery-beyond-our-borders.pdf. Accessed 5th January 2023.

16. Sandwell, R. et al. 2018. Stronger together: Midwifery twinning between Tanzania and Canada. Globalization and Health 14(1). https://dx.doi.org/10.1186/s12992-018-0442-x.

17. Taniguchi, H. et al. 2021. Twinning Project between the Japanese Midwives Association and Mongolian Midwives Association for Organisational Strengthening as Shown by MACAT. Research Square.

18. United Nations. 2015. Transforming our World: The 2030 Agenda for Sustainable Development. New York: Available from: https://sdgs.un.org/publications/transforming-our-world-2030-agenda-sustainable-development-17981 . Accessed 5th January 2023.

